# Identification of SARS-CoV-2-specific immune alterations in acutely ill patients

**DOI:** 10.1101/2020.12.21.20248642

**Authors:** Rose-Marie Rébillard, Marc Charabati, Camille Grasmuck, Abdelali Filali-Mouhim, Olivier Tastet, Nathalie Brassard, Audrey Daigneault, Lyne Bourbonnière, Renaud Balthazard, Ana Carmena Moratalla, Yves Carpentier Solorio, Negar Farzam-kia, Antoine Philippe Fournier, Elizabeth Gowing, Hélène Jamann, Florent Lemaître, Victoria Hannah Mamane, Karine Thai, Jean-François Cailhier, Nicolas Chomont, Andrés Finzi, Michaël Chassé, Madeleine Durand, Nathalie Arbour, Daniel E. Kaufmann, Alexandre Prat, Catherine Larochelle

## Abstract

Dysregulated immune profiles have been described in symptomatic SARS-CoV-2-infected patients. Whether the reported immune alterations are specific to SARS-CoV-2 infection or also triggered by other acute illnesses remains unclear. We performed flow cytometry analysis on fresh peripheral blood from a consecutive cohort of i) patients hospitalized with acute SARS-CoV-2 infection; ii) patients of comparable age/sex hospitalized for other acute disease (SARS-CoV-2 negative); and iii) healthy controls. Using both data-driven and hypothesis-driven analyses, we found several dysregulations in immune cell subsets (e.g. decreased proportion of T cells) that are similarly associated with acute SARS-CoV-2 infection and non-COVID-19 related acute illnesses. In contrast, we identified specific differences in myeloid and lymphocyte subsets that are associated with SARS-CoV-2 status (e.g. elevated proportion of ICAM-1^+^ mature/activated neutrophils, ALCAM^+^ monocytes, and CD38^+^CD8^+^ T cells). A subset of SARS-CoV-2-specific immune alterations correlated with disease severity, disease outcome at 30 days and mortality. Our data provides novel understanding of the immune dysregulation that are specifically associated with SARS-CoV-2 infection among acute care hospitalized patients. Our study lays the foundation for the development of specific biomarkers to stratify SARS-CoV-2^+^ patients at risk of unfavorable outcome and uncover novel candidate molecules to investigate from a therapeutic perspective.

## Introduction

The severe acute respiratory syndrome coronavirus 2 (SARS-CoV-2) that causes the coronavirus-19-disease (COVID-19) has now infected millions of people worldwide and caused the death of over 1 million patients (WHO operational update on November 6^th^ 2020). The spectrum of clinical manifestations in SARS-CoV-2-infected patients (SARS-CoV-2^+^) ranges from asymptomatic to severe acute respiratory distress syndrome (ARDS) and multiple organ involvement (1). Increasing evidence supports that the immune reaction plays a central role in COVID-19 severity and outcome. Therefore, it is essential to understand immune responses generated in COVID-19 to stratify patients at higher risk of unfavorable outcome and identify novel therapeutic targets. Several risk factors for negative clinical outcome have been identified (2, 3) but the underlying mechanisms of effective vs. impaired/deleterious immune responses to SARS-CoV-2 remain unclear.

Age is the greatest risk factor for COVID-19 severity and mortality (2-6). Obesity and cardiovascular comorbidities such as high blood pressure and diabetes also significantly increase the risk of severe clinical course in SARS-CoV-2-infected individuals (3, 4, 6, 7). Notably, each of these risk factors –age, obesity, diabetes and cardiovascular comorbidities– is associated with alterations in immune responses and a state of chronic low-grade inflammation (8-10) that could contribute to the elevated morbidity and mortality in COVID-19, as in other infectious conditions (11-13). Dysregulations of the immune profile in SARS-CoV-2-infected hospitalized patients (COVID-19) include elevated levels of circulating IL-6 and decreased peripheral lymphocytes/neutrophils ratios, which are predictive of worse clinical outcome (7, 14, 15). Accumulating evidence suggest that both hyperactivation and exhaustion of different T and B cell subsets characterize COVID-19 (7, 14, 16). A state of activation of CD4^+^ T cells with an increased CD4^+^/CD8^+^ ratio has been recently linked to disease severity (14, 17). Whether the reported immune alterations are specific to SARS-CoV-2 infection or rather commonly triggered by a range of acute illnesses, especially in older subjects with pre-existing medical conditions, remains unknown. We performed a detailed characterization of circulating innate and adaptive immune cells from 50 SARS-CoV-2^+^ patients (47 hospitalized for COVID-19, three asymptomatic nosocomial infection) in comparison with 22 patients similar in regards to sex and age hospitalized for other acute illnesses (SARS-CoV-2^neg^) and 49 healthy controls (HC). We found a SARS-CoV-2-specific immune profile that could identify relevant therapeutic targets related to severity and outcome in COVID-19 patients.

## Results

### SARS-CoV-2^+^ and SARS-CoV-2^neg^ groups of hospitalized patients have similar prevalence of comorbidities, severity of disease and outcome at 30 days

To investigate SARS-CoV-2-triggered alterations in the peripheral immune system, we performed an extensive immune profiling of whole blood obtained from hospitalized patients and HC. Inclusion criteria for hospitalized participants included a nasopharyngeal swab SARS-CoV-2 PCR test conducted because of symptoms compatible with COVID-19, or because they were considered at risk of acute SARS-CoV-2 infection. Patients were classified as SARS-Cov-2^+^ patients or SARS-CoV-2^neg^ patients based on the test results. Mean age and sex ratio were similar in SARS-CoV-2^+^ and SARS-CoV-2^neg^ hospitalized patients’ groups, while HC were younger and included a higher proportion of women (Table 1). No significant differences were observed in numbers of comorbidities, history of cancer or organ transplant between the SARS-CoV-2^+^ and SARS-CoV-2^neg^ hospitalized patients (Table 1). Most hospitalized patients presented at least one comorbidity regardless of coronavirus status. A greater proportion of SARS-CoV-2^+^ subjects had comorbidities associated with risk for severe COVID-19 (obesity, chronic renal insufficiency and type 2 diabetes) compared to SARS-CoV-2^neg^ patients, without reaching significance (Table 1). In contrast, respiratory and hepatic conditions were more frequent among SARS-CoV-2^neg^ patients (not significant), probably reflecting the patient population in our hospital.

**Table 1:** Baseline characteristics of population

| Baseline clinical characteristics | Healthy controls<br>(n = 49) | Hospitalized<br>SARS-CoV-2<sup>neg</sup><br>(n = 22) | Hospitalized<br>SARS-CoV-2<sup>+</sup><br>(n = 50) | Statistical analysis<br>(SARS-CoV-2<sup>neg</sup> vs<br>SARS-CoV2<sup>+</sup>) |
| --- | --- | --- | --- | --- |
| Age, mean years (SD) | 38.7 (11.9) | 58.4 (17.2) | 59.5 (14.6) | # p = 0.9347 |
| Male sex, n (%) | 20 (40.8) | 15 (68.2) | 30 (60.0) | * p = 0.6022 |
| <60y.o., n (%) | 49/49 (100) | 9/22 (40.9) | 20/50 (40) | * p > 0.9999 |
| *Co-morbidities and past medical history* |  |  |  |  |
| BMI over 25.0, n (%) | ND | 13/22 (59.1) | 38/46 (82.6) | * p = 0.0698 |
| Chronic renal insufficiency, n (%) | ND | 1/22 (4.5) | 11/50 (22.0) | * p = 0.0904 |
| Cardiac condition, n (%) | ND | 5/22 (22.7) | 9/50 (18.0) | * p = 0.7485 |
| Respiratory condition, n (%) | ND | 7/22 (31.8) | 7/50 (14.0) | * p = 0.1073 |
| Hepatic condition, n (%) | ND | 3/22 (13.6) | 1/50 (2.0) | * p = 0.0820 |
| High blood pressure, n (%) | ND | 11/22 (50.0) | 33/50 (66.0) | * p = 0.2939 |
| Dyslipidemia, n (%) | ND | 7/22 (31.8) | 22/50 (44.0) | * p = 0.4362 |
| Diabetes type 1, n (%) | ND | 0/22 (0) | 1/50 (2.0) | * p > 0.9999 |
| Diabetes type 2, n (%) | ND | 6/22 (27.2) | 24/50 (48.0) | * p = 0.1242 |
| Co-morbidities total,<br>n (excluding obesity) (%) | ND | 0: 6/22 (27.2)<br>1-2: 9/22 (40.9)<br>≥ 3: 7/22 (31.8) | 0: 11/50 (22.0)<br>1-2: 17/50 (34.0)<br>≥ 3: 22/50 (44.0) | ‡ p = 0.6235 |
| Organ transplant, n (%) | ND | 3/22 (13.6) | 4/50 (8.0) | * p = 0.6678 |
| Active cancer, n (%) | ND | 3/22 (13.6) | 3/50 (6.0) | * p = 0.3612 |
| Cancer history, n (%) | ND | 4/22 (18.2) | 4/50 (8.0) | * p = 0.2372 |
| Active infection except COVID-19, n (%) | ND | 10/22 (45.5) | 16/50 (32.0) | * p = 0.2979 |

SD: standard deviation; n: number of patients in specified category; ND: not determined; y.o.: years old; + Fisher's exact test; # Mann-Whitney U test; ‡ Chi-squared test.

SARS-CoV-2^+^ and SARS-CoV-2^neg^ hospitalized patients’ groups exhibited comparable outcome at 30 days, mortality rate, and need for invasive ventilation during hospitalization (Table 2). Acute renal insufficiency was more frequent in SARS-CoV-2^+^ but did not reach significance. As previously reported, SARS-CoV-2 infection was associated with a high incidence of respiratory insufficiency and encephalopathy (delirium) in hospitalized patients (2, 3, 18). The total burden of medical complications during hospitalization and the average duration of invasive ventilation were elevated in SARS-CoV-2^+^ patients (Table 2). Of note, at time of baseline blood sampling, both groups of patients showed a similar proportion of intensive care unit (ICU) hospitalization and number of days in hospital (Supplemental Table 1). Association analyses among clinical parameters (Figure 1A) demonstrated that our two patient groups presented the expected relationships between age, comorbidities (past medical history: PMH) and medical complications. Moreover, risk of death correlated with mechanical ventilation, respiratory insufficiency, and other medical complications. We also observed the expected association of SARS-CoV-2 status with typical COVID-19 symptoms at admission, as well as with delirium and respiratory insufficiency. Age, disease severity, and unfavorable outcome were associated but the overlap was only partial (Table 2).

**Table 2:** Clinical data related to hospitalization in SARS-CoV-2^neg^ and SARS-CoV-2^+^ patients

| Clinical data-Hospitalization | SARS-CoV-2<sup>neg</sup><br>(n = 22) | SARS-CoV-2<sup>+</sup><br>(n = 50) | Statistical analysis |
| --- | --- | --- | --- |
| *Outcome and severity* |  |  |  |
| Symptomatic COVID-19, n (%) | NA | 47/50 (94) | NA |
| Severe disease (high flow or invasive ventilation), n (%) | 14/22 (63.6) | 21/50 (42) | * p = 0.1255 |
| <60y.o. in mild-moderate disease, n (%) | 3/8 (37.5) | 12/29 (41.4) | * p > 0.9999 |
| <60y.o. in severe disease, n (%) | 6/14 (42.9) | 8/21 (38.1) | * p > 0.9999 |
| Thirty-day outcome (NIH 8 point-scale), mean (SD) | 6.3 (2.3) | 5.7 (2.5) | * p = 0.3724 |
| Thirty-day outcome: ambulatory (7-8 on NIH 8 point-scale), n (%) | 16/22 (72.7) | 27/50 (54.0) | * p = 0.1930 |
| Thirty-day outcome: unfavorable (1-4 on NIH 8 point-scale), n (%) | 5/22 (22.7) | 14/50 (28) | * p = 0.7751 |
| <60y.o. in thirty-day outcome: unfavorable, n (%) | 2/5 (40) | 3/14 (21.4) | * p = 0.5696 |
| Thirty-day outcome: deceased, n (%) | 2/22 (9.1) | 3/50 (6.0) | * p = 0.6379 |
| Sixty-day outcome: deceased, n (%) | 2/22 (9.1) | 7/50 (14.0) | * p = 0.7125 |
| Invasive ventilation during hospitalization, n (%) | 10/22 (45.5) | 21/50 (42.0) | * p = 0.8017 |
| Total duration of invasive ventilation, median day (range) | 4.5 (1-20) | 24.5 (8-57) | # p < 0.0001 |
| *Medical complications during hospitalization* |  |  |  |
| Thrombo-embolic event excluding admission for NSTEMI, n (%) | 2/22 (9.1) | 7/50 (14.0) | * p = 0.7125 |
| Acute renal insufficiency, n (%) | 3/22 (13.6) | 18/50 (36.0) | * p = 0.0896 |
| Cardiac event, n (%) | 5/22 (22.7) | 12/50 (24.0) | * p > 0.9999 |
| Respiratory insufficiency, n (%) | 9/22 (40.9) | 31/50 (62.0) | * p = 0.1251 |
| Elevated liver enzymes, n (%) | 8/22 (36.4) | 23/50 (46.0) | * p = 0.6061 |
| Delirium, n (%) | 2/22 (9.1) | 20/50 (40.0) | * p = 0.0115 |
| Complications total, n (%) | 0: 5/22 (22.7)<br>1-2: 14/22 (63.6)<br>≥ 3: 3/22 (13.6) | 0: 12/50 (24.0)<br>1-2: 14/50 (28.0)<br>≥ 3: 24/50 (48.0) | ‡ p = 0.0074 |

n: number of patients in specified category; y.o.: years old; NA: not applicable
+ Fisher's exact test; # Mann-Whitney U test; ‡ Chi-squared test

**Figure 1.**
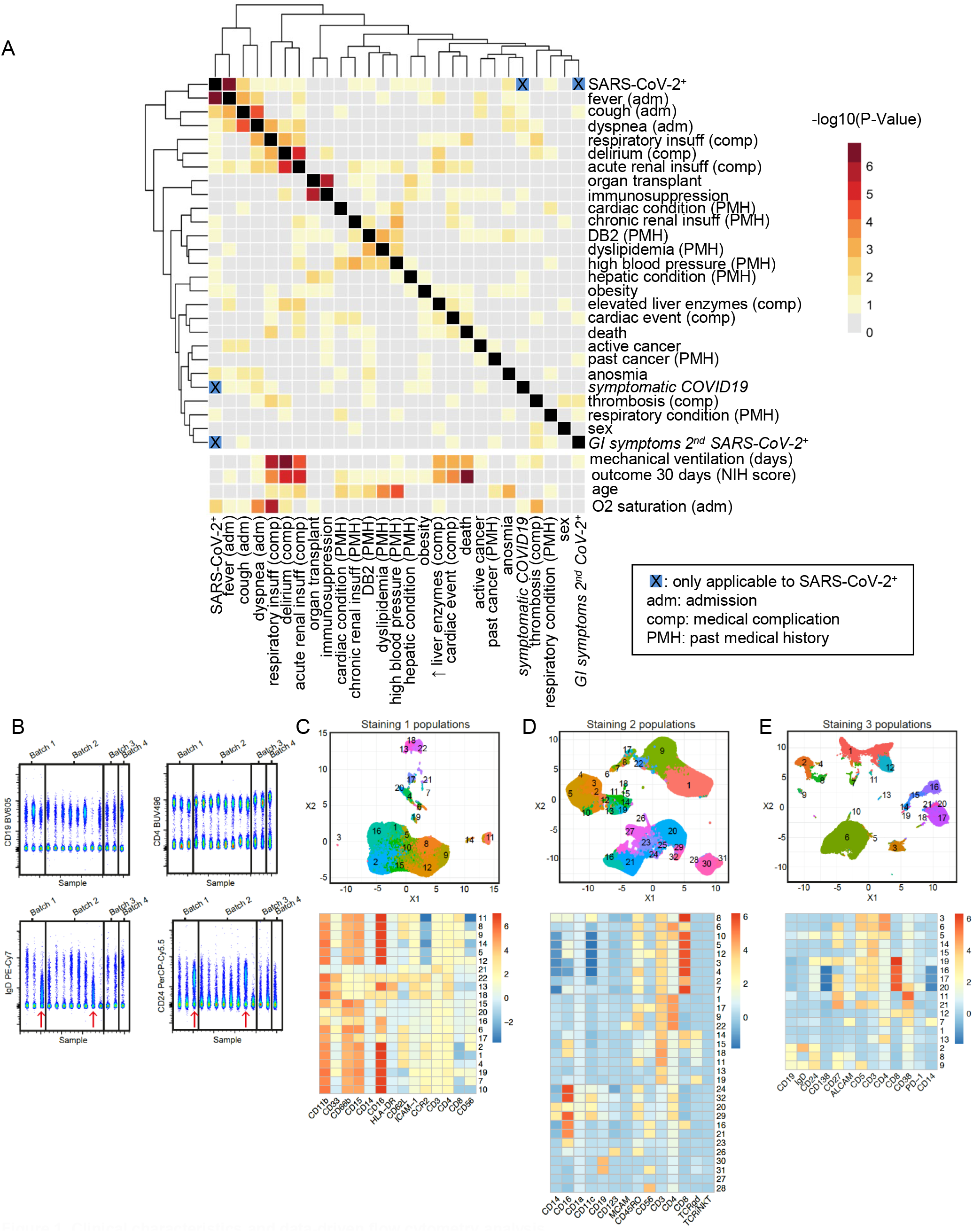
Clinical characteristics and data-driven flow cytometry analysis. (**A**) Association among clinical parameters in hospitalized patients as illustrated by heatmap and hierarchical clustering of the -log10 (*P-*value). Fisher exact test for association of binary variables (upper part), and Wilcoxon rank sum test for association between binary and continuous variables (lower part). Blue squares indicate that only SARS-CoV-2^+^ patients were considered for this parameter. (**B**) Representative dot plots of multiple samples acquired in four different batches for CD19-BV605, CD4-BUV496, IgD-PE-Cy7 and CD24-PerCP-Cy5.5. Red arrows identify samples from the same individual acquired in two different batches. (**C-D-E**) Representation of FlowSOM populations on UMAP projection axis and their corresponding heatmap showing geometric mean fluorescence of the different markers for the different subpopulations (unbiased analysis). Cellular markers are indicated on the x-axis and population number on the y-axis of heatmaps.

### Dysregulation of peripheral immune cell subsets in SARS-CoV-2^+^ and SARS-CoV-2^neg^ hospitalized patients

To compare the immune profiles of SARS-CoV-2^+^ and SARS-CoV-2^neg^ hospitalized patients and HC, we performed three complementary flow cytometry staining panels on fresh whole blood samples and assessed the frequency of different subpopulations of lymphocytes, monocytes, dendritic cells and granulocytes. Our quality control analysis on the first 20 samples acquired in four batches over two weeks ruled out significant batch effects and validated the capture of reproducible inter-individual variation (example IgD detection, Figure 1B). We used two analysis approaches: i) a data-driven approach, using the PhenoGraph (19) and FlowSOM (20) algorithms to cluster similar cells in an unbiased manner (Figure 1C-E), and ii) a hypothesis-driven analysis with conventional gating strategy. We focused our analysis on cell subsets representing at least 1% of major cell subsets. Using the data-driven approach, we identified 21 cell clusters in the monocyte and neutrophil gate (gating according to granularity and size excluding lymphocytes, 14 markers) for the first staining panel (S1) (Figure 1C and Supplemental Figure 1A), 31 cell clusters in the monocyte and lymphocyte gates for the dendritic cell/NK cell-oriented panel (S2, 14 markers, gating excluding granulocytes) (Figure 1D and Supplemental Figure 1A) and 18 cell clusters in the monocyte and lymphocyte gate for the lymphocyte-oriented panel (S3, 13 markers, gating excluding granulocytes) (Figure 1E and Supplemental Figure 1A). The distribution of clustered populations as illustrated by the Uniform Manifold Approximation and Projection (UMAP) algorithm shows distinct populations and relative abundance of these subsets in blood samples.

We compared the abundance of these 70 identified cell clusters in the three donor groups: HC, SARS-CoV-2^+^ hospitalized patients and SARS-CoV-2^neg^ hospitalized patients. Only populations with at least one significant (adjusted p value < 0.05) difference between two groups were considered. We identified 13 immune subpopulations that were specifically increased or decreased in samples from SARS-CoV-2^+^ hospitalized patients compared to both HC and SARS-CoV-2^neg^ hospitalized patients (Figure 2A, populations identified in green) and 16 subpopulations that were similarly altered in both groups of hospitalized patients compared to HC (Figure 2A, populations identified in blue). The distribution of some of these subpopulations showing significant differences between our groups are illustrated (Figure 2B and C). We observed alterations in the abundance of peripheral immune cell subpopulations (e.g. T cells, B cells, myeloid/monocytes and neutrophils) compared to HC in the acutely ill groups regardless of SARS-CoV-2 status, for example a subset of CD19^+^ B cells (S2_Pop30) (Figure 2B). In contrast, the abundance of specific immune cell subsets was elevated or reduced in SARS-CoV-2^+^ compared to SARS-CoV-2^neg^ hospitalized patients, for example a subset of CD27^+^CD8^+^ T cells (S3_Pop17) (Figure 2C).

**Figure 2.**
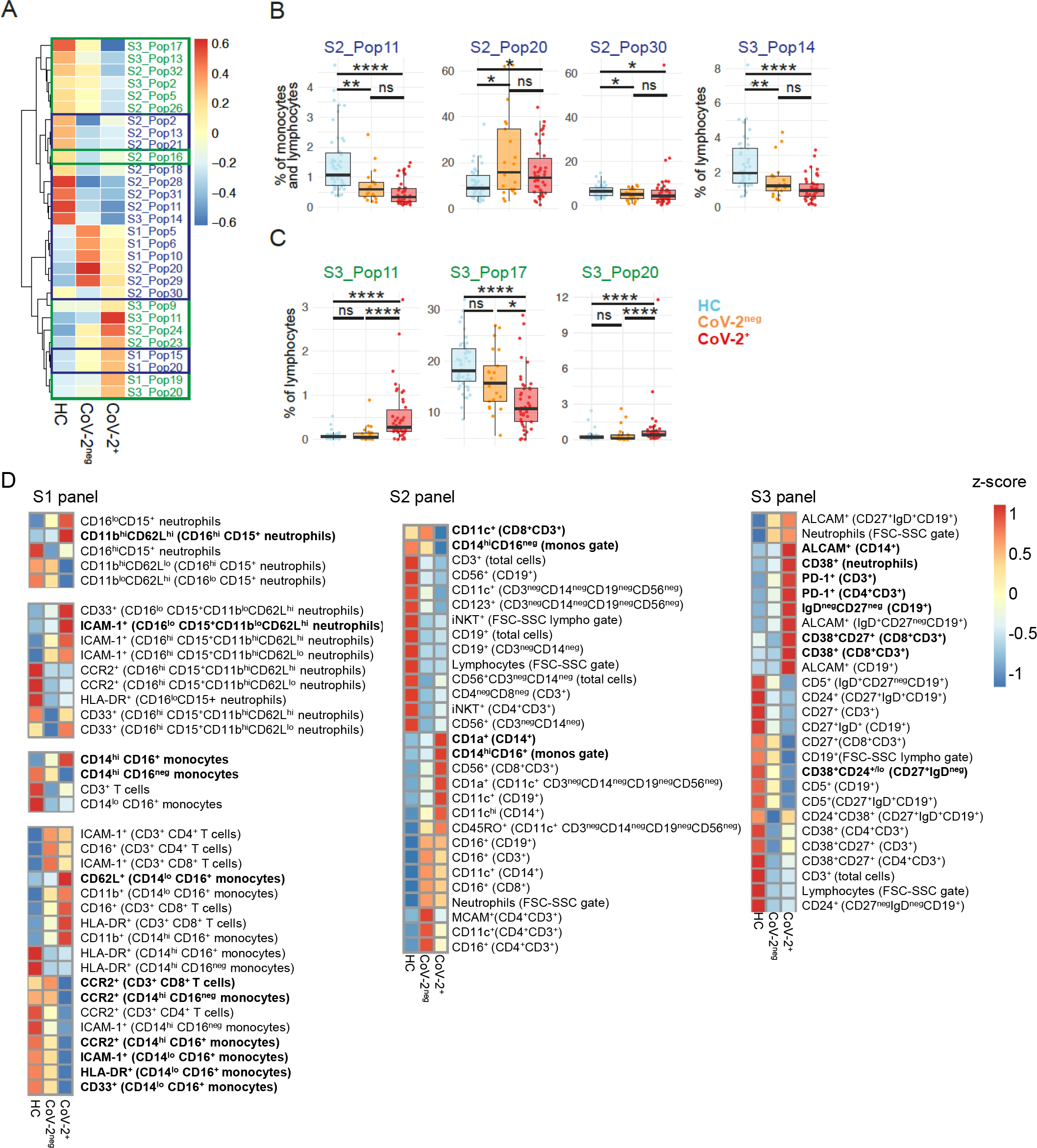
Common and distinct alterations in immune cell populations in SARS-CoV-2^+^ and SARS-CoV-2^neg^ hospitalized patients. (**A-C**) Data-driven analysis. (**A**) Heatmap showing the median frequencies of immune cell populations differentially regulated in SARS-CoV-2^+^ patients (CoV-2^+^), SARS-CoV-2^neg^ (CoV-2^neg^) and healthy control (HC) samples. (**B-C**) Box and Whisker plots showing frequencies of dysregulated immune cell populations in (**B**) CoV-2^+^ (red) compared to CoV-2^neg^ (yellow) and HC (blue) and in (**C**) hospitalized patients (both CoV-2^+^ and CoV-2^neg^) compared to HC (blue). HC, *n* = 49; CoV-2^neg^, *n* = 21; CoV-2^+^, *n* = 42. Kruskal-Wallis test followed by a Dunn’s post hoc-test for multiple pairwise comparisons. \**P* < 0.05; \*\**P* < 0.01; \*\*\*\**P* < 0.0001. (**D**) Hypothesis-driven (conventional) analysis. Heatmaps showing the median frequency of immune cell populations identified as significantly altered (adjusted *P* < 0.05) in CoV-2^+^ (*n* = 50) compared to HC (*n* = 49) and/or CoV-2^neg^ (*n* = 22) in stain 1 (S1 panel), stain 2 (S2 panel) and (stain 3 (S3 panel). Kruskal-Wallis test followed by a Dunn’s post hoc-test for multiple pairwise comparisons. Nominal *P*-values were adjusted for multiple testing within each stain, false discovery rate significance threshold set at 0.05. Color scale legends the z-score, across the groups, of sub-population median frequencies. Populations significantly altered (*P* < 0.05) in SARS-CoV-2^+^ compared to HC and to SARS-CoV-2^neg^ samples are in **bold**.

To corroborate these observations, we used a conventional approach to analyse the different subsets of granulocytes, myeloid/monocytes and lymphocytes. Only those with significant differences (adjusted p value < 0.05) according to status (HC or SARS-CoV-2^neg^ or SARS-CoV-2^+^) are presented in Figure 2D. In line with the results of the unbiased analysis, we found several immune cell populations that differed significantly from HC in hospitalized patients but are associated with acute illness rather than being specific for COVID-19. Of note, the differences observed according to status (HC or SARS-CoV-2^neg^ or SARS-CoV-2^+^) in general immune cell populations (e.g. lymphocytes) were consistent across the different panels (Figure 2D). Importantly, we were able to identify significant and specific alterations in samples from SARS-CoV-2^+^ compared to both HC and SARS-CoV-2^neg^ hospitalized patients, for example upregulation of PD-1 on T cells (particularly CD4^+^ T cells) (Figure 2D populations indicated in bold).

### Neutrophilia and lymphopenia predict outcome but are associated with severity of illness and age rather than specific to SARS-CoV-2 infection

Previous studies have identified lymphopenia and neutrophilia as predictive of worse clinical outcome in SARS-CoV-2 infection (8,15,16). Compared to HC, we detected a significantly increased proportion of neutrophils, and a corresponding decreased proportion of lymphocytes (CD3^+^ T cells and CD19^+^ B cells), in samples from hospitalized patients (Figure 2 and 3). However, these changes were not specific for SARS-CoV-2 infection and were observed in both hospitalized groups. As age represents an important factor shaping the immune system as well as one of the greatest risk factors for COVID-19 severity and mortality (2-6), we also compared these immune populations in HC and younger patients (<60 y.o.), as well as in older patients (≥ 60 y.o.). We observed that lymphopenia and neutrophilia were most striking in older patients (Figure 3A) and in relation to the severity of the medical condition, independently of SARS-CoV-2 status or other acute illness status (Figure 3B). While not specific for SARS-CoV-2 infection, neutrophilia and lymphopenia, as well as a low proportion of CD3^+^ but not CD19^+^ cells, were in addition strongly tied to outcome at 30 days (Figure 3C). Within the neutrophil compartment, we observed a statistically significant shift from CD16^hi^CD15^+^ to CD16^lo^CD15^+^ neutrophils in SARS-CoV-2^+^ and SARS-CoV-2^neg^ patients compared with HCs (Figure 2D and 3A and Supplemental Figure 3), but no difference when comparing SARS-CoV-2^+^ with SARS-CoV-2^neg^ patients and no association with disease severity (Figure 3B), which demonstrates that this shift was not specific to COVID-19. Within the CD3^+^ T cell compartment, the CD4^+^/CD8^+^ ratio increased with older age as expected but was not different between groups of similar age, while CD3^+^ γδT cells and iNKT cells were reduced in SARS-CoV-2^+^ samples compared to HC but not different from SARS-CoV-2^neg^ samples in hospitalized patients nor associated with disease severity (Figure 3A-B).

**Figure 3.**
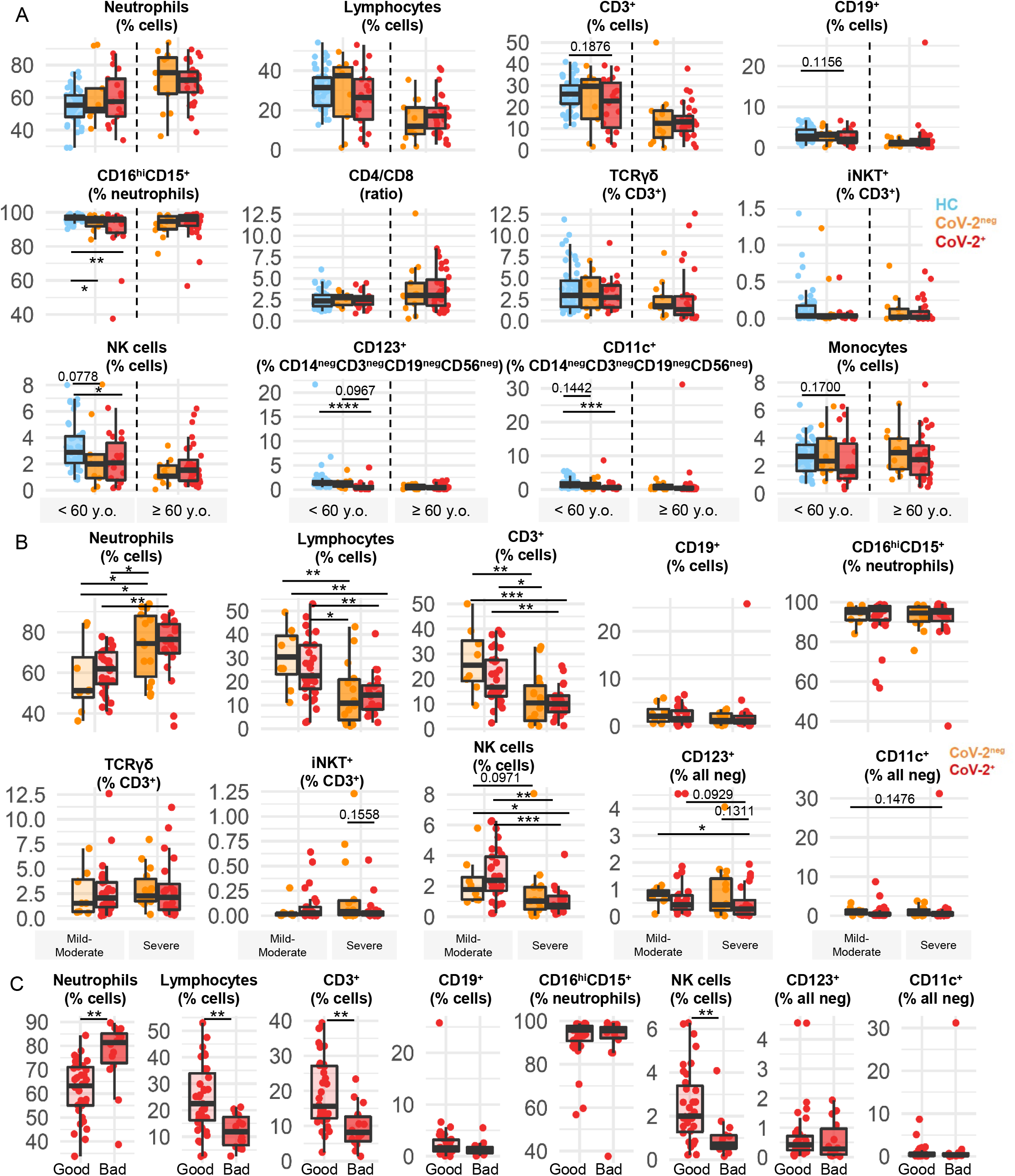
Alterations in immune cell populations commonly observed in SARS-CoV-2^+^ and SARS-CoV-2^neg^ hospitalized patients. (**A-C**) Frequencies of different subsets of immune cell populations in peripheral blood (conventional analysis) from SARS-CoV-2^+^ (CoV-2^+^, red) and SARS-CoV-2^neg^ (CoV-2^neg^, yellow) hospitalized patients and healthy controls (HC, blue) (**A**) according to age group (HC < 60 y.o. n = 49; CoV-2^neg^ hospitalized < 60 y.o. *n* = 9; ≥ 60 y.o. *n* = 13; CoV-2^+^: < 60 y.o. *n* = 20; ≥ 60 y.o. n = 30), (**B**) according to disease severity in hospitalized patients (CoV-2^neg^ mild/moderate disease *n* = 8, severe disease *n* = 14; CoV-2^+^ mild/moderate disease *n* = 29, severe disease *n* = 21), and (**C**) according to clinical outcome at 30 days in SARS-CoV-2^+^ patients (NIH score 5-8, *n* = 36) vs. (NIH score 1-4, *n* = 14). Mann-Whitney *U* test (for *n* = 2 categories) and Kruskal-Wallis test (for *n >* 2 categories) followed by Dunn’s post-hoc test for the multiple pairwise comparisons were used. Each dot represents one donor. \**P* < 0.05; \*\**P* < 0.01; \*\*\**P* < 0.001; \*\*\*\**P* < 0.0001.

The frequency of NK cells was reduced in both groups of hospitalized patients compared to HC (Figure 2D), especially in individuals with severe medical condition (COVID-19 or other acute illness), and was significantly lower in SARS-CoV-2^+^ patients showing a worse clinical outcome at 30 days (Figure 3A-C). Finally, as previously reported (21), we observed a decreased frequency of CD123^+^ and CD11c^+^ cells among the CD3^neg^CD19^neg^CD14^neg^CD56^neg^ cells (presumably pDCs and cDCs) in the peripheral blood of SARS-CoV-2^+^ patients but this was not significantly different in SARS-CoV-2^neg^ acutely ill patients. The frequency of monocytes was similar in HC, SARS-CoV-2^neg^ and SARS-CoV-2^+^ patients (Fig. 3A) but HLA-DR expression on CD14^+^ monocytes was significantly reduced in both groups of hospitalized patients as compared to HC (Fig. 2D), indicating a possible monocyte exhaustion, a phenomenon typically observed following acute intense monocyte inflammation as seen for example in sepsis (22). Altogether, our data confirm previous reports linking neutrophilia and lymphopenia with severe COVID-19 and worse outcome; however, we demonstrate that these changes are associated with age and severity of the medical condition rather than being specific for SARS-CoV-2 infection.

### SARS-CoV-2 infection is associated with a significantly higher proportion of circulating pro-myelocyte and mature neutrophils but not activated neutrophils

As significant neutrophilia was observed in both SARS-CoV-2^neg^ and SARS-CoV-2^+^ patients compared to HC, we investigated whether specific neutrophil subsets within the granulocytes’ gate (CD14^neg^ cells in the neutrophil FSC-SSC gate) were specifically altered in SARS-CoV-2^+^ patients. The proportion of promyelocytes (CD38^+^), precursors of granulocytes, was significantly higher in SARS-CoV-2^+^ patients compared to SARS-CoV-2^neg^ patients and HC (Figure 2D); this increased percentage of promyelocytes was however not significantly different according to severity, and did not distinguish outcome at 30 days (Figure 4A-C). In parallel, the frequency of mature neutrophils (CD16^hi^CD15^+^CD11b^hi^CD62L^hi^) was specifically increased in the peripheral blood in SARS-CoV-2^+^ patients (Figure 2D), while that of activated neutrophils (CD16^hi^CD15^+^CD11b^hi^CD62L^lo^) was lower, but these subsets were not associated with severe COVID-19 (Figure 4B and Supplemental Figure 3B). Our results suggest that specific stages of neutrophils were over-represented in peripheral blood of SARS-CoV-2^+^ patients, while most circulating activated neutrophils might have already infiltrated organs (e.g. lungs), died of over-stimulation (NETosis) following acute infection and/or were immunosuppressed as previously reported (23).

**Figure 4.**
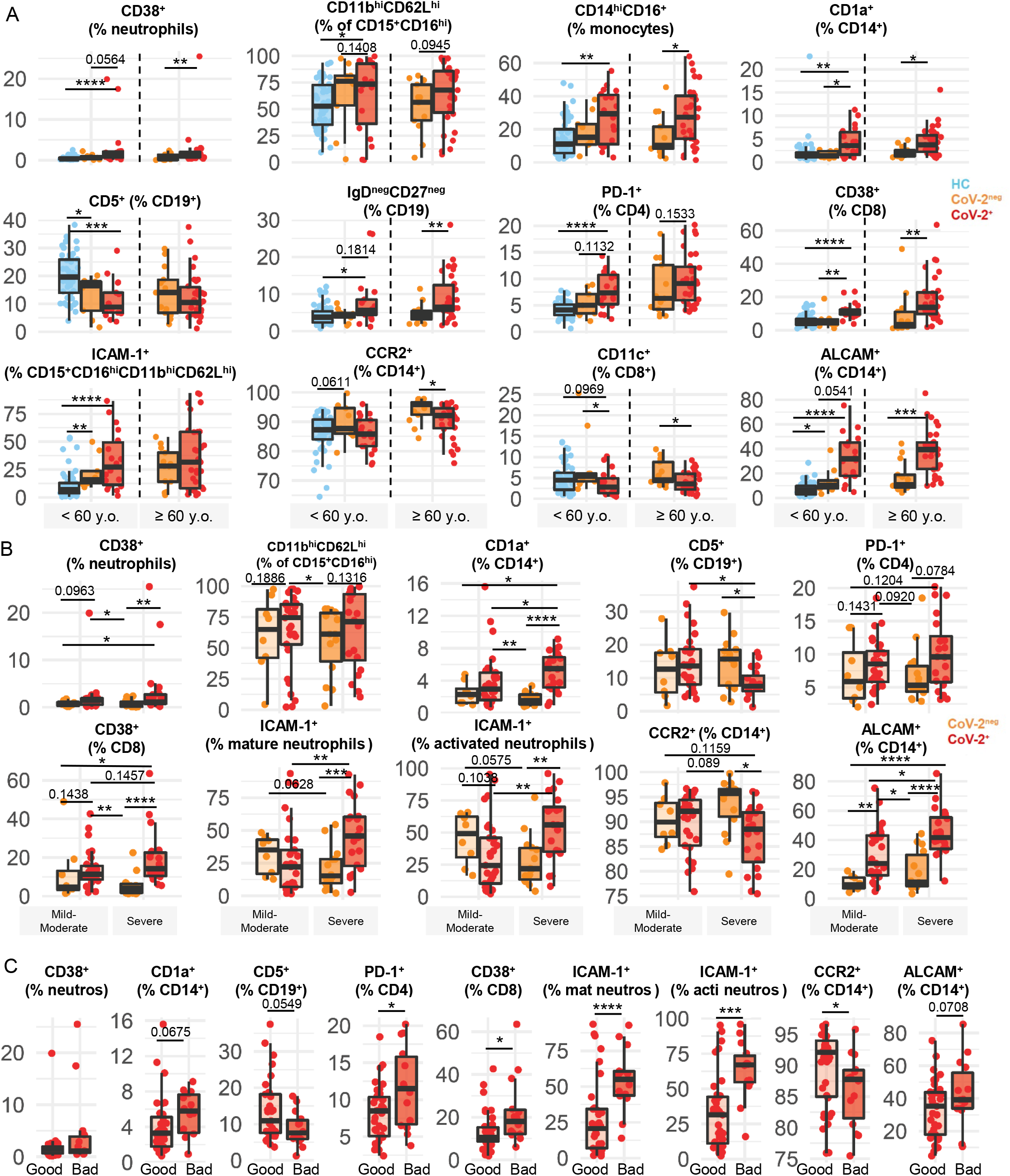
Alterations in immune cell populations distinguishing SARS-CoV-2^+^ from SARS-CoV-2^neg^ hospitalized patients. (**A-C**) Frequencies of different subsets of immune cell populations in peripheral blood (conventional analysis) from SARS-CoV-2^+^ (CoV-2^+^, red) and SARS-CoV-2^neg^ (CoV-2^neg^, yellow) hospitalized patients and healthy controls (HC, blue) (**A**) according to age groups (HC < 60 y.o. *n* = 49; CoV-2^neg^ hospitalized < 60 y.o. *n* = 9, ≥ 60 y.o. *n* = 13; CoV-2^+^: < 60 y.o. *n* = 20, ≥ 60 y.o. *n* = 30), (**B**) according to disease severity in hospitalized patients (CoV-2^neg^ mild/moderate disease *n* = 8, severe disease *n* = 14; CoV-2^+^ mild/moderate disease *n* = 29, severe disease *n* = 21), and (**C**) according to clinical outcome at 30 days in SARS-CoV-2^+^ patients (NIH score 5-8, *n* = 36) vs. (NIH score 1-4, *n* = 14). Mann-Whitney *U* test (for *n* = 2 categories) and Kruskal-Wallis test (for *n >* 2 categories) followed by Dunn’s post-hoc test for the multiple pairwise comparisons were used. Each dot represents one donor. \**P* < 0.05; \*\**P* < 0.01; \*\*\**P* < 0.001; \*\*\*\**P* < 0.0001.

### Expression of MHC-related molecule CD1a on monocytes is associated with SARS-CoV-2 status, severity and outcome

We also investigated whether specific subpopulations of antigen presenting cells (APC) were altered in patients as these could secrete multiple inflammatory mediators (cytokines, myeloperoxidases, reactive oxidative species, etc.) and present antigen to T cells. Within the monocyte compartment, monocytes bearing CD16 (FCγIIIR) are associated with secretion of pro-inflammatory cytokine TNF, antigen-presenting capacity and tissue homing (24). The proportion of classical CD14^hi^CD16^neg^ monocytes was specifically reduced in SARS-CoV-2^+^ patients compared to HC and SARS-CoV-2^neg^ patients; in contrast, the frequency of intermediate CD14^hi^CD16^+^ monocytes was specifically augmented in SARS-CoV-2^+^ patients (Figure 2D and 4A), but was not significantly associated with severity (Supplemental Figure 3B) or outcome (data not shown). Notably, the proportion of CD14^+^ monocytes expressing CD1a, which is implicated in lipid antigens presentation to T cells and is induced on CD14^+^ monocytes exposed to IL-4 and GM-CSF (25), was also increased in SARS-CoV-2^+^ patients compared to both HC and SARS-CoV-2^neg^ patients regardless of age (Figure 2D and 4A). Moreover, CD1a^+^CD14^+^ monocytes were more prevalent in SARS-CoV-2^+^ patients exhibiting severe symptoms and in SARS-CoV-2^+^ patients experiencing unfavorable outcome at 30 days. Overall, our results suggest that in response to SARS-CoV-2 infection, specific subsets of peripheral blood antigen presenting cells are altered.

### B cells subsets are altered in SARS-CoV-2 infection

B cell-depleting therapies are associated with a higher risk of severe COVID-19 course (26) and several publications support the contribution of neutralizing antibodies to the immune response in SARS-CoV-2 infection (27-30) As we detected a reduction in peripheral blood B cells in hospitalized patients compared to HCs, we assessed the frequency of specific B cell subpopulations in our cohort using CD5, as well as CD27 and IgD as subset markers. We found that the proportion of B cells expressing CD5 was significantly reduced in SARS-CoV-2^neg^ and SARS-CoV-2^+^ patients compared to HC (Figure 2D), but this reduction was more pronounced in SARS-CoV-2^+^ patients, especially in patients experiencing a severe clinical course or showing an unfavorable clinical outcome at 30 days (Figure 4A-C). Notably, CD5^+^ B cells play a crucial role in innate immunity (31) and include natural polyreactive antibody-producing cells (32) and IL-10-producing regulatory cells (33). Analysis of B cells according to their expression of IgD and CD27 revealed a significantly higher proportion of IgD^neg^CD27^neg^ B cells in SARS-CoV-2^+^ patients compared to HC and to SARS-CoV-2^neg^ patients (Figure 2D); this increase was more pronounced in older patients (Figure 4A) but did not differ according to severity (Supplemental Figure 3B) or outcome (data not shown). This IgD^neg^CD27^neg^ B cell subset is expanded in the peripheral blood of patients with inflammatory diseases, viral infections and upon aging (34-37). In contrast, the other subpopulations (CD27^+^IgD^neg^, CD27^+^IgD^+^, CD27^neg^IgD^+^) were either similarly prevalent in all three groups or comparable between SARS-CoV-2^neg^ and SARS-CoV-2^+^ patients (Figure 2D and Supplemental Figure 3A). Further classification of subsets of naïve, unswitched memory, switched memory and double negative B cells according to CD24 and CD38 expression did not reveal expression patterns specific to SARS-CoV-2 infection except for a reduction in the abundance of pre-switched/resting memory B cells (CD27^+^IgD^neg^CD24^+^CD38^+/lo^) that was more pronounced in older SARS-CoV-2^+^ patients (Figure 2D and Supplemental Figure 3A) but was not associated with disease severity (p = 0.41) or outcome (p = 0.45). Overall, our results demonstrate alterations of specific B cell subpopulations (reduced CD5^+^ and increased IgD^neg^CD27^neg^) in SARS-CoV-2^+^ patients compared to the other groups, suggesting a shift towards a pro-inflammatory B cell profile.

### Activated CD38^+^CD8^+^ T cells and PD-1^+^CD4^+^ T cells characterize SARS-CoV-2^+^ patients and are associated with outcome

We investigated whether multiple markers (PD-1, CD38, CD27, CD56) triggered on activated or memory T cell subpopulations and associated with antiviral responses are altered in SARS-CoV2^+^ patients (Figure 2D). CD38 has been linked to T cell functions such as protection from cell death (38) and high suppressive activity by regulatory T cells, and to disease progression in HIV patients (38). We found that CD38 expression was strongly up-regulated on CD8^+^ T cells in SARS-CoV-2^+^ patients compared to SARS-CoV-2^neg^ patients and HC (Figure 2D) regardless of age (Figure 4A). Levels tended to be higher in severe COVID-19 compared to mild/moderate SARS-CoV-2 infection (Figure 4B). Importantly, the percentage of CD38^+^CD8^+^ T cells was significantly higher in the worse outcome group at 30 days (Figure 4C). Our results suggest that an elevated frequency of CD38^+^CD8^+^ T cells represents a biomarker for a protracted course of severe COVID-19 still requiring supplemental oxygenation at 30 days. We observed a significant reduction in the proportion of CD4^+^ T cells expressing CD38 in both SARS-CoV-2^neg^ and SARS-CoV-2^+^ patients compared to HC (Figure 2D) without any significant association with age, severity or outcome (Supplemental Figure 3).

PD-1 is an immune checkpoint molecule associated with T cell activation and exhaustion (39). The proportion of PD-1-expressing CD3^+^ T cells and more specifically CD4^+^ T cells was elevated in both older and younger SARS-CoV-2^+^ patients compared to SARS-CoV-2^neg^ hospitalized patients and HC (Figure 2D and 4A). PD-1 expression was not associated with disease severity among SARS-CoV-2^+^ samples (Figure 4B) but the percentage of PD-1^+^CD4^+^ T cells was significantly elevated in SARS-CoV-2^+^ patients demonstrating an unfavorable outcome at 30 days (Figure 4C). Expression of PD-1 on CD8^+^ T cells was higher in SARS-CoV-2^+^ patients compared to HC but this was almost exclusively observed in older patients and did not significantly differ from SARS-CoV-2^neg^ hospitalized patients (Supplemental Figure 3A). Finally, the percentage of CD8^+^ T cells expressing CD27 and CD56 were respectively reduced and increased compared to HC, and showed a similar trend compared to SARS-CoV-2^neg^ patients, but were not associated with severity (Figure 2D and Supplemental Figure 3A-B). Overall, our results show that CD38 on CD8^+^ T cells and PD-1 on CD4^+^ T cells were specifically altered in SARS-CoV-2^+^ patients and associated with outcome at 30 days.

### Up-regulation of specific trafficking molecules distinguishes SARS-CoV-2 infection, COVID-19 severity and clinical outcome

Diapedesis of circulating leukocytes is a crucial inflammatory step in immune responses that, when out of control, can cause serious collateral damage (40, 41). To determine whether COVID-19 affects the trafficking potential of circulating leukocytes, we assessed a selection of key trafficking molecules (ICAM-1, ALCAM, CCR2, CD11c) on the different immune cell populations. In granulocytes we observed that the proportion of mature and activated neutrophils expressing ICAM-1 (42) is increased in hospitalized patients compared to HC and more so in SARS-CoV-2^+^ patients (Figure 2D and 4A and Supplemental Figure 3), especially those exhibiting a severe disease and experiencing an unfavorable outcome at 30 days (Figure 4B-C). These data suggest that ICAM-1^+^ neutrophils, which have been associated with dissemination of inflammation through reverse transendothelial migration, cellular aggregation and effector function (43-45), could participate to fuel the pro-inflammatory cascade observed in COVID-19.

Strikingly, the percentage of monocytes (CD14^+^) expressing the cell adhesion molecule ALCAM was strongly up-regulated in SARS-CoV-2^+^ patients compared to both HC and SARS-CoV-2^neg^ patients (Figure 2D and 4A). This increased frequency was found in younger and older patients and was significantly higher in patients with severe COVID-19 and elevated in patients who showed unfavorable outcome at 30 days (Figure 4B-C). ALCAM is associated with leukocyte transendothelial migration and with the stabilization of the immune synapse (46, 47). This suggest that monocytes from SARS-CoV-2^+^ patients could be poised to activate lymphocytes and interact with the endothelium, thus participating in the general inflammatory processes.

Inversely CCR2, a chemokine receptor implicated in recruitment of monocytes to pulmonary alveolar tissue (48), was expressed by a reduced percentage of monocytes (CD14^+^) in SARS-CoV-2^+^ patients compared to SARS-CoV-2^neg^ patients (Figure 2D and 4A). This reduction was not associated with disease severity but was significantly more pronounced in SARS-CoV-2^+^ patients with an unfavorable outcome at 30 days (Fig. 4B-C). As the bronchoalveolar fluid from COVID-19 patients has been shown to contain abundant inflammatory monocyte-derived macrophages and CCR2 ligand CCL2 (49), the reduced proportion of CCR2^+^ monocytes in the periphery could result from preferential transmigration of these subsets in the inflamed lungs, or simply reflect the reduction in classical monocytes which also express CCR2.

Among T cells, CCR2 expression was also reduced in SARS-CoV-2^+^ patients (Figure 2D) but did not significantly correlate with severity or outcome (good vs. bad outcome: CD4^+^CCR2^+^ 14.42% ± 1.11 vs 11.80% ± 1.72 p = 0.24; CD8^+^CCR2^+^ 8.53% ± 1.07 vs. 7.09% ± 1.32, p = 0.71). The frequency of CD8^+^ T cells bearing integrin β2 CD11c was significantly lower in the blood of SARS-CoV-2^+^ patients compared to SARS-CoV-2^neg^ (Figure 2D and 4A), especially in severe cases (Supplemental Figure 3B), but did not correlate with outcome among SARS-CoV-2^+^ patients (p = 0.33). Notably, this T cell subset has anti-viral properties and can accumulate in the lungs during infection (50), and can suppress pathogenic T cells in autoimmunity (51). The proportion of B cells expressing CD11c or ALCAM followed a similar pattern (Figure 2D), with a specific up-regulation of both markers on B cells from SARS-CoV-2^+^ especially in patients over 60 y.o. compared to HC and SARS-CoV-2^neg^ patients; however, there was no significant difference between mild/moderate and severe COVID-19 symptoms (Supplemental Figure 3B) or according to outcome among SARS-CoV-2^+^ patients.

### Multivariate analysis confirms independent associations of identified immune cell populations with SARS-CoV-2 status

Using random forest class prediction analysis (52), we explored whether the immune cell populations identified by the univariate non-parametric Kruskal-Wallis tests as being associated to SARS-CoV-2 status are also independently associated to status in a multivariate model. We confirmed that frequency of ICAM-1^+^ activated neutrophils, CCR2^+^, CD1a^+^ and ALCAM^+^ monocytes, CD38^+^CD8^+^ T cells and PD-1^+^CD4^+^ T cells, were all among the top 30 important predictive features, as measured by the Gini importance index (Supplemental Figure 4)(52), to discern between HC and hospitalized patients. However, CD5^+^ B cells were not found to be of high importance to distinguish between HC and SARS-CoV-2^+^ patients in this multivariate analysis. Of note, HC and SARS-CoV-2^+^ patients but not SARS-CoV-2^neg^ patients were classified with a high accuracy. Such inaccuracy to classify SARS-CoV-2^neg^ patients is probably due to the lower number of patients and the greater heterogeneity of diseases in this group.

### COVID-19 mortality is associated with higher frequency of ICAM-1^+^ neutrophils, ALCAM^+^ monocytes and CD38^+^CD8^+^ T cells

We next determined whether the immune subpopulations we identified as dysregulated in hospitalized patients (24 populations) are associated with selected clinical parameters (Figure 5A). Sex, obesity and days since onset of symptoms did not show strong correlations with most immune parameters. While not specific to SARS-CoV-2^+^ patients, neutrophilia and lymphopenia, especially low CD3^+^ T cells count, were associated with medical complications, invasive ventilation and mortality in SARS-CoV-2^+^ patients. In addition, we found that most immune subpopulations specifically altered in SARS-CoV-2^+^ patients, and linked with severity and outcome, were similarly associated with medical complications, invasive ventilation and mortality (Figure 5A and Table 3). When looking specifically among severe COVID-19 patients requiring invasive ventilation, those who did not survive at 60 days had a significantly higher proportion of ICAM-1^+^ cells among activated and mature neutrophils (Figure 5B). In addition, deceased patients showed a higher proportion of monocytes expressing ALCAM^+^ but a trend towards a reduced proportion expressing CCR2. Finally, a higher proportion of CD38^+^CD8^+^ T cells (Figure 5A-B) was observed in deceased severe COVID-19 patients compared to survivors. The other SARS-CoV-2^+^ specific dysregulated immune populations (PD-1^+^CD4^+^ T cells, CD5^+^ on B cells, CD1a^+^ monocytes) were not significantly altered in deceased compared to survivors among severe COVID-19 patients (Table 3).

**Table 3:** Summary of identified alterations in subsets of immune cells according to status, severity, outcome and mortality

| Immune cell population \ Association | Status (CoV-2<sup>+</sup> vs. CoV-2<sup>neg</sup>) | Severity COVID-19 (CoV-2<sup>+</sup>) | Outcome at 30 days (CoV-2<sup>+</sup>) | Mortality at 60 days (severe CoV-2<sup>+</sup>) |
| --- | --- | --- | --- | --- |
| Neutrophils | ↔ | ↑ | ↑ | ↗ |
| Lymphocytes | ↔ | ↓ | ↓ | ↘ |
| CD3<sup>+</sup> T lymphocytes | ↔ | ↓ | ↓ | ↘ |
| CD19<sup>+</sup> B lymphocytes | ↔ | ↔ | ↔ | ↔ |
| Monocytes | ↔ | ↓ | ↘ | ↓ |
| NK cells | ↔ | ↓ | ↓ | ↘ |
| iNKT | ↔ | ↔ | ↔ | ↔ |
| TCRγδ | ↔ | ↔ | ↔ | ↘ |
| PD-1<sup>+</sup> CD4<sup>+</sup> T lymphocytes | ↑ | ↔ | ↑ | ↔ |
| CD11c<sup>+</sup> CD8<sup>+</sup> T lymphocytes | ↓ | ↘ | ↔ | ↔ |
| CD38<sup>+</sup> CD8<sup>+</sup> T lymphocytes | ↑ | ↗ | ↑ | ↑ |
| CD27<sup>neg</sup>IgD<sup>neg</sup> B lymphocytes | ↑ | ↔ | ↔ | ↔ |
| CD5<sup>+</sup>CD19<sup>+</sup> | ↘ | ↓ | ↘ | ↔ |
| CD16<sup>hi</sup>CD15<sup>+</sup> neutrophils | ↔ | ↔ | ↔ | ↔ |
| CD123<sup>+</sup> in CD14<sup>neg</sup> myeloid | ↔ | ↘ | ↔ | ↘ |
| CD11c<sup>+</sup> in CD14<sup>neg</sup> myeloid | ↔ | ↔ | ↔ | ↔ |
| CD38<sup>+</sup> promyelocytes | ↑ | ↔ | ↔ | ↑ |
| Mature neutrophils | ↑ | ↔ | ↘ | ↔ |
| ICAM-1<sup>+</sup> in mature and activated neutrophils | ↗ | ↑ | ↑ | ↑ |
| CD14<sup>hi</sup>CD16<sup>+</sup> monocytes | ↑ | ↔ | ↔ | ↓ |
| ALCAM<sup>+</sup> monocytes | ↑ | ↑ | ↗ | ↗ |
| CCR2<sup>+</sup> monocytes | ↓ | ↔ | ↓ | ↘ |
| CD1a<sup>+</sup> monocytes | ↑ | ↑ | ↗ | ↔ |

CoV-2<sup>+</sup>: SARS-CoV-2<sup>+</sup>; CoV-2<sup>neg</sup>: SARS-CoV-2<sup>neg</sup>

**Figure 5.**
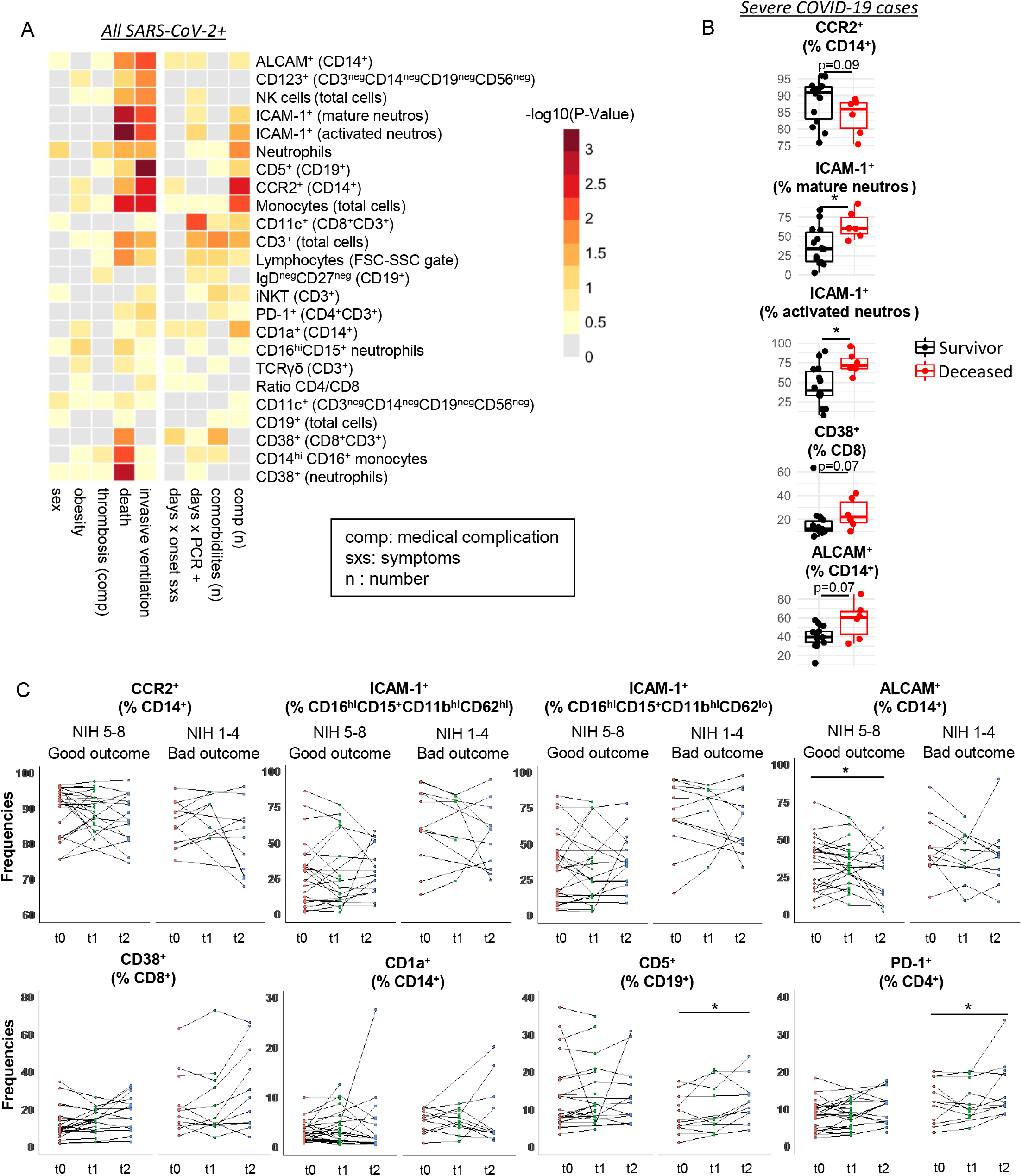
Association between clinical parameters and longitudinal analysis of immune cell populations altered in SARS-CoV-2^+^ patients. (**A**) Association between immune cell subsets and clinical parameters in SARS-CoV-2^+^ patients as illustrated by heatmap and hierarchical clustering of the -log10 (*P*-value) analyzed by Mann-Whitney *U* test for categorical clinical parameters and by Spearman’s correlation for continuous clinical parameters. (**B**) Proportions of different immune cell subsets among severe SARS-CoV-2^+^ patients (severe COVID-19) according to survival at 60 days. Mann-Whitney *U* test were used. (**C**) Changes over time in frequencies of selected populations identified as specific to SARS-CoV-2^+^, between baseline (t0), 24-72h (t1) and 4-7 days (t2). Generalized estimating equations analysis. Each dot represents one patient. \**P* < 0.05.

### Longitudinal analysis reveals increasing expression of CD38 and PD-1 on T cells in patients with unfavorable outcome

We were able to obtain follow-up samples at 24h-72h (t1) and at 4-8 days (t2) after the baseline sample (t0) in a subset of SARS-CoV-2^+^ patients. Using a semi-parametric approach, Generalized Estimation Equations(53), we examined the trend over time in selected populations identified as specific to COVID-19. We did not observe a significant trend for ICAM-1^+^ neutrophils, or CD1a^+^ or CCR2^+^ monocytes, but we found that expression of ALCAM on monocytes was consistently and significantly lower over time in SARS-CoV-2^+^ showing a favorable outcome, but not in those showing an unfavorable outcome, at 30 days (Figure 5B). On the contrary, while the patients with a good outcome at 30 days did not show a significant change in the proportion of their B cells expressing CD5 over time, patients with a bad outcome almost all presented an increase at follow-up, starting from lower levels at first sampling. The proportion of CD38^+^CD8^+^ T cells doubled over time in patients with COVID-19 regardless of outcome, therefore the increased frequency in patients with a bad outcome remained present at follow-up. Finally, while levels were not significantly different over time in SARS-CoV-2^+^ patients displaying a favorable outcome at 30 days, expression of PD-1 on CD4^+^ T significantly increased over time in most patients experiencing a bad outcome at 30 days and were strongly up-regulated at follow-up in 3/4 patients who did not survive.

## Discussion

Understanding the specific immune responses associated with SARS-CoV-2 infection is paramount to the quest for targeted therapy. Many groups have reported immune dysregulations in circulating leukocytes from SARS-CoV-2 infected patients compared to healthy, often younger, subjects (14-16, 54, 55). Most of them however did not investigate whether these changes were specific to SARS-CoV-2 infection or represent nonspecific remodeling of the immune system in response to stress inherent to acute illness. Indeed, other critical non-infectious medical conditions are associated with immune perturbations (56). To discriminate the specific impact of SARS-CoV-2 infection on the immune system, we took a broader approach and included hospitalized SARS-CoV-2^neg^ patients that are acutely ill and comparable to SARS-CoV-2^+^ patients in regards to age and sex and performed a whole blood flow cytometry analysis. We found multiple immune dysregulations in hospitalized patients compared to healthy subjects (Figure 2). Nevertheless, we established specific differences in myeloid (e.g. elevated proportion of ALCAM^+^ monocytes) and lymphocyte (e.g. increased percentage of CD38^+^CD8^+^ T cells) subpopulations in SARS-CoV-2 infected patients compared to SARS-CoV-2^neg^ patients (Figure 4A). Finally, a subset of these immune alterations correlated with disease severity and outcome (Figure 3B-C, 4B-C, 5A-B and Table 3). Overall, our study provides the groundwork to develop specific peripheral blood biomarkers to stratify SARS-CoV-2^+^ patients at risk of unfavorable outcome and identify novel candidate molecules as potential therapeutic targets.

We observed changes in the proportions of key white blood cell subsets: neutrophils and lymphocytes. Indeed, we detected lymphopenia and neutrophilia in hospitalized patients regardless of their SARS-CoV-2 status; the most severe perturbations were found in older patients and associated with disease severity as well as disease outcome (Figure 3). Although other groups have linked severe SARS-CoV-2 infection with lymphopenia (57), our results as well as those from others (58) support the notion that lymphopenia is in fact commonly observed in critically ill patients, especially in older patients, and not related to the SARS-CoV-2 infection (56, 59). Nevertheless, we found alterations in both innate and adaptive immune cell subpopulations that were specifically associated with SARS-CoV-2 infection.

We observed elevated proportions of pro-inflammatory innate immune cell subsets. Notably, the frequency of CD38^+^ neutrophils was increased in SARS-CoV-2^+^ patients (Figure 4). CD38 expression during neutrophil activation has been shown to play an essential role for appropriate control of pathogen infection and cell migration in affected organs (38). SARS-CoV-2^+^ patients had a higher proportion of CD14^hi^CD16^+^ monocytes than SARS-CoV-2^neg^ patients. This monocyte subset shows robust reactive oxygen species (ROS) and TNF production, strongly promotes proliferation and antigen presentation to T cells (24). In addition, the frequency of CD1a^+^CD14^+^ monocytes was higher in SARS-CoV-2^+^ compared to SARS-CoV-2^neg^ patients and such alteration was associated with worse clinical outcome (Figure 4C). CD1a^+^CD14^+^ monocytes can activate and induce differentiation of pro-inflammatory IFN-γ^+^ CD4^+^ T cells (60). We found a depletion of CD5^+^ B cells, which comprise B-1a cells and play a crucial role in innate immunity, in our two groups of hospitalized patients, although this depletion was more pronounced in SARS-CoV-2^+^ patients and most striking in SARS-CoV-2^+^ patients with unfavorable outcome (Figure 4C). Such depletion has been described in sepsis (61) and SARS-CoV-2 infection (55, 62). CD5^+^ B cells can secrete inhibitory cytokines (63) and natural IgM as an early response to pathogen invasion (31), and promote tissue homeostasis and facilitate clearance of apoptotic cells (31). B-1a cells regulate neutrophil lung infiltration, inhibit the production of myeloperoxidase and modulate macrophage responses in ARDS (64). As recently reviewed, further investigations will be necessary to elucidate whether B-1a depletion in SARS-CoV-2^+^ patients facilitates aggressive inflammatory response and promotes lung damage (65).

We identified altered expression of cell adhesion molecules on peripheral immune cells from SARS-CoV-2^+^ patients and an association of such changes with the outcome, including mortality, among patients with severe COVID-19 (Figure 4, 5B and Table 3). We observed elevated ICAM-1^+^ on mature and activated neutrophils in SARS-CoV-2^+^ patients compared to their SARS-CoV-2^neg^ counterparts. Others have suggested that ICAM-1 is acquired by neutrophils as they exit the inflamed tissue to re-enter the circulation (66). This process might contribute to the clearance of neutrophils from the site of injury (67) but can also participate to dissemination of inflammation and promotion of distant organ damage (42, 68). Furthermore, ICAM-1^+^ neutrophils are associated with the formation of neutrophil extracellular traps (NET) (69, 70), which is a network of DNA, histone and protein associated with thrombosis. Notably, elevated ICAM^+^ neutrophil subset was associated with the burden of medical complications (Figure 5A). We further found that ALCAM was present on an elevated proportion of monocytes, and to a lesser extent B cells, in SARS-CoV-2^+^ individuals compared to other groups (Figure 2D and 4A). This elevated proportion of ALCAM^+^ monocytes was associated with severe and fatal COVID-19 and persisted in cases with a bad outcome at 30 days. ALCAM is associated with transmigration of monocytes across pulmonary endothelium (71) and with T cell activation; it could therefore represent a novel relevant therapeutic target in COVID-19.

We detected significant and persistent alterations in the T cell compartment in SARS-CoV2^+^ patients compared to other groups. CD38^+^CD8^+^ T cells were more prominent in SARS-CoV-2^+^ patients compared to other groups especially in patients who experienced an unfavorable outcome and did not survive. It is possible that CD38^+^CD8^+^ T cells represent overactivated, potentially exhausted and less efficient T cells in viral control, as suggested by HIV studies (72, 73). We observed an elevated proportion of T cells carrying PD-1 in both groups of hospitalized patients with higher levels in elderly patients (Figure 2). PD-1 expression on CD4^+^ was more pronounced in SARS-CoV-2^+^ patients and in patients who required a prolonged stay in the ICU (unfavorable outcome at 30 days). Whereas it is possible that PD-1 upregulation is a physiological attempt to limit immunopathology in the setting of immune hyperactivation and plays a positive role, taken together, our findings raise the question about the therapeutic potential of PD-1/PD-L1 pathway modulation to reverse immune exhaustion in SARS-CoV-2^+^ patients, specifically in elderly patients (74-76). Notably, elderly septic patients –particularly those with unfavorable prognosis– exhibit a prolonged lymphopenia, a preferential reduction of CD4^+^ T cells as well as an elevated expression of PD-1 on CD4^+^ and CD8^+^ T cells (77-80). In animal models of sepsis, blockade of the PD-1/PD-L1 pathway restored T cell function and was associated with a decrease of the pathogen burden and better survival (76, 81-83). Whether such approach will be beneficial in the context of COVID-19 will soon be determined by an ongoing phase II trial for treatment of SARS-CoV-2 infected patients with an anti-PD-1 monoclonal antibody (ClinicalTrials.gov Identifier: NCT04356508).

From a clinical point of view, the similarities uncovered between the immune dysregulation in SARS-CoV-2^+^ and SARS-CoV-2^neg^ patients suggest that therapeutics targeting general non-specific inflammatory processes could be tested in multiple severe acute illness when lymphopenia and neutrophilia are observed, especially in older patients. Some therapies successfully used in sepsis or other ICU care settings could be applied to COVID-19, as exemplified by the use of steroids in severe COVID-19 (84).

Our study has identified new biomarkers specifically associated with unfavorable outcome in COVID-19 patients. These surface markers could represent relevant potential therapeutic targets to explore in future studies, in particular PD-1 on CD4^+^ T cells, and ICAM-1 and ALCAM on neutrophils and antigen presenting cells (B cells, monocytes). Finally, our longitudinal investigation has revealed markers that could predict worse outcome and correlate with medical complications and mortality (Figure 5A and Table 3). We believe our experimental approach, whole blood staining and flow cytometry analysis, could be easily used in hospital diagnostic biochemistry laboratories to follow patients overtime and could complement other clinical follow-ups.

## Methods

### Cohort

Fifty SARS-CoV-2 positive patients and twenty-two SARS-CoV-2 negative patients admitted to the University of Montreal hospital center (CHUM) between April 7^th^ and May 7^th^ 2020, as well as 49 healthy controls from the CHUM personnel, were prospectively recruited to the study and included in the Biobanque Québécoise de la COVID-19 (BQC19). None of the patients included received an experimental treatment for COVID-19 (e.g. hydroxychloroquine, remdesevir, anti-IL-6) prior to peripheral blood sampling. One SARS-CoV-2^+^ patient had to be excluded from analysis related to outcome and survival for participation after baseline sampling in an experimental pharmacological assay. SARS-CoV-2^+^ and SARS-CoV-2^neg^ status was determined by PCR (repeated once when negative) among hospitalized patients. The absence of IgM or IgG antibodies against SARS-CoV-2 in the serum of SARS-CoV-2^neg^ hospitalized patients and of healthy controls was used to confirm the status of the donors. When possible follow-up samples were obtained at 24h-72h (t1) and at 4-8 days (t2) after the baseline sample (t0) for SARS-CoV-2^+^ patients. A follow-up sample at 24-72h was obtained from 32 SARS-CoV-2^+^ subjects, at 4-7 days from 28 subjects and at both timepoints for 21 subjects. For some subanalyses, patients were stratified according to age: younger <60 y.o. vs. older ≥ 60 y.o. Hospitalization in the ICU was used to define other severe acute illness in SARS-CoV-2^neg^ patients, and the need for high-flow or invasive ventilation in SARS-CoV-2^+^ patients to define severe COVID-19. In our cohort 14/22 patients were classified as other severe acute illness and 21/50 as severe COVID-19. The 8-point NIH Ordinal Severity scale (ranging from 1 = death to 8 = discharged at home with no required care) was used to describe the outcome at 30 days. A score of 1-4 on the NIH scale was used to define an unfavorable or bad outcome, while a score of 5 to 8 was used to define a favorable or good outcome at 30 days. Mortality up to 60 days after sampling was considered when indicated. Medical charts were reviewed by two independent physicians to collect clinical information.

### Flow cytometry

Blood was obtained through venous puncture and collected in tubes containing trisodium citrate, citric acid and dextrose (ACD) tubes. 150 ul whole blood was used for each staining (3 stainings/sample). Red blood cells were lysed using a 15min incubation with BD PharmLyse solution (#555899) at room temperature according to the manufacturer’s instructions. After subsequent washing, samples were blocked for 15min with 50 uL of mouse IgG (Invitrogen #10400C) diluted 1:10. Three antibody panels were used, details of which are summarized in Supplemental Tables 2-4. BD Brilliant Buffer (#566349) was added to all antibody cocktails to minimize staining artifacts. Following 20 minutes incubation at 4°C, cells were washed and fixed with BD Cytofix solution (#554655) for 20 minutes. Samples were acquired on a BD FACSFortessa. BD FACSDiva™ CS&T research beads (#655051) were acquired biweekly to ensure the stability of the cytometer. SPHERO supra rainbow fluorescent particles (Spherotech #SRCP-35-2A) were recorded prior to sample acquisition to assess laser instabilities. Onecomp eBeads (Invitrogen #01-1111-42) and UltraComp eBeads (Invitrogen #01-3333-42) were used for compensation and manual correction for each antibody panel was performed before analysis and applied to all samples.

### Flow cytometry analysis

Flow cytometric data analysis was performed using FlowJo (version 10.6.2). For both biased and unbiased analysis, doublets were excluded, and events were pre-gated on the FSC-A vs. SSC-A biplot to define granulocytic, monocytic and lymphocytic cell lineages based on size and granularity. Compensation matrices were calculated and applied. Data-driven analysis was performed on select cell lineages, depending on the antibodies included in each panel: analysis of the first panel was performed on the events within the monocytic gate and the granulocytic gate, analysis of the second panel on the events within the monocytic gate and the lymphocytic gate, and analysis of the third panel on the events within the lymphocytic gate. Samples were then downsampled to an equal size and one large concatenated file was generated for each panel. Using the R packages flowCore 2.0.1 and FlowSOM 1.20.0 in R version 4.0.1, we applied the FlowSOM algorithm(20) on these concatenated files to create a FlowSOM map for each panel. The modal value of clusters, as determined by the PhenoGraph clustering algorithm(19) in FlowJo on multiple random samples, was used to determine the number of clusters to input into FlowSOM. A heatmap of each panel’s geometric mean fluorescence intensity (MFI) for each cellular cluster was generated with the R package pheatmap 1.0.12. UMAPs were generated using the R package UMAP 0.2.6.0.

### Statistics

Analysis of frequency of populations and their association with SARS-CoV-2 status, severity and outcome at 30 days were performed by Mann-Whitney U test (for factors with 2 categories) or by Kruskal-Wallis test (for factors with a number of categories > 2) followed by Dunn’s post-hoc test. All tests were performed two-sided, using a nominal significance threshold of p < 0.05. For identification of populations of interest (Figure 2A and 2D), nominal p-values for Kruskal-Wallis tests were adjusted for Multiple testing (adjusted p value) within each stain separately using the method of Benjamin and Hochberg (85), which controls the false discovery rate at a significance threshold we set at 0.05. Association between clinical categorical variables was assessed by fisher exact test. Correlation between continuous features were quantified by the Spearman rank correlation. To assess frequency of specific immune cell populations over time, longitudinal analysis was performed using generalized estimation equation as implemented in the R package gee using exchangeable correlation structure and including time as the fixed effect variable and patient ID as the random variable.

Multivariate prediction of the SARS-CoV-2^+^, SARS-CoV-2^neg^ and HC status, using as candidate predictor variables the whole set of the immune subpopulations, was performed using a random forest (52) classification models as implemented in the randomForest 4.6-14 R package using 1000 random trees and the default “mtry” (number of variables randomly sampled and tested in each node) parameter. A misclassification error for each status was calculated as the out-of-bag error by testing on the samples which were not randomly drawn at each tree generation. All statistical analyses were performed using the statistical package R version 4.0.1.

### Study approval

This study was approved by the CRCHUM ethic committee in accordance with the Declaration of Helsinki (IRB protocols 19.387 and 19.389). Informed consent was obtained for each patient and is detailed elsewhere(86).

## Data Availability

n/a

## Author contributions

C.L., A.P., D.E.K., N.A., R.M.R, M.Charabati and C.G. designed the study and wrote the manuscript; the order of co-first authors was determined by flipping a coin; C.L., A.P., D.E.K., and N.A. supervised the study; A.F, J.F.C and N.C. provided scientific input for designing the study and writing the manuscript; R.B., Y.C.S., N.F., A.P.F., E.G., H.J., F.L., V.A.M., A.C.M, N.B., A.D., L.B. and K.T. performed experiments; C.L., A.F.M., O.T., R.M.R, M.Charabati and C.G. performed analysis; M.Chassé and M.D. recruited patients for the study.

## Funding

This study was funded by a grant from the CRCHUM foundation and by grant VR2-173203 from the Canada’s COVID-19 Immunity Task Force (CITF), in collaboration with the Canadian Institutes of Health Research (CIHR). The Biobanque Québécoise de la COVID-19 (BQC-19) is supported by the Fonds de recherche Québec-Santé (FRQS), Génome Québec and the Public Health Agency of Canada. D.E.K is a FRQS Merit Research Scholar. N.C., M.D. and C.L. receive a salary award from the FRQS. A.Finzi. and A.P. hold a Canada Research Chair.

We gratefully acknowledge Dominique Gauchat and Philippe St-Onge from the flow cytometry platform from CRCHUM for excellent technical guidance and support.

