## Supplemental for "Identification of SARS-CoV-2-specific immune alterations in acutely ill patients"

### Supplemental material

Supplemental Table 1: Clinical data related to the day of sampling (baseline sample)

| **Sampling** | **SARS-CoV-2$^{neg}$ (n = 22)** | **SARS-CoV-2$^{+}$ (n = 50)** | **Statistical analysis** |
|---|---|---|---|
| Time from onset of COVID-19 symptoms to baseline sampling, median day (range) | NA | 12.0 (3-23) | NA |
| Time from first documented positive SARS-CoV-2 PCR to baseline sampling, median day (range) | NA | 6.5 (2-23) | NA |
| Time from hospital admission to baseline sampling, median day (range) | 3.5 (1-22) | 4.5 (1-61) | $^{\#}$ p = 0.4209 |
| $O_2$ requirements at time of baseline sampling, n (%) | 13/22 none (59.1)<br>4/22 low flow (18.2)<br>5/22 high flow/invasive (22.7) | 21/50 none (42.0)<br>8/50 low flow (16.0)<br>21/50 high flow/invasive (42.0) | $^{‡}$ p = 0.3418 |
| In ICU at time of baseline sampling n (%) | 10/22 (45.5) | 23/50 (46.0) | $^{+}$ p > 0.9999 |

n: number of patients in specified category; NA: not applicable
+ Fisher's exact test; # Mann-Whitney U test; ‡ Chi-squared test

Supplemental Table 2: Granulocyte & monocyte-oriented panel (S1)

| Fluorochrome | Antigen | Manufacturer/Catalogue # | Clone |
|---|---|---|---|
| FITC | CD66b | BD-555724 | G10F5 |
| PE | CD33 | BioLegend-303404 | WN53 |
| APC | ICAM-1 | BD-559771 | HA58 |
| v450 | CD15 | BD-561584 | HI98 |
| AF700 | HLA-DR | BD-560743 | G46-6 |
| PE-CF594 | CD16 | BD-562320 | 3G8 |
| PerCP-Cy5.5 | CCR2 | BD-357203 | K036C2 |
| Pe-Cy7 | CD11b | eBioscience-25-0118-42 | ICRF44 |
| BV605 | CD56 | BioLegend-318334 | HCD56 |
| BV785 | CD62L | Biolegend-304830 | DREG-56 |
| BV711 | CD8 | BD-563677 | RPA-T8 |
| BUV395 | CD3 | BD-563546 | UCHT1 |
| BUV496 | CD4 | BD-612936 | SK3 |
| BUV737 | CD14 | BD-564444 | M5E2 |

Supplemental Table 3: NK Cell/Dendritic Cell-oriented Panel (S2)

| Fluorochrome | Antigen | Manufacturer/Catalogue # | Clone |
|---|---|---|---|
| FITC | CD1a | BD-555806 | HI149 |
| PE | TCR iNKT | BioLegned-342904 | 6B11 |
| APC | MCAM | Miltenyi -130-120-701 | 541-10B2 |
| BV421 | CD123 | BD-562517 | 9F5 |
| APC-R700 | CD11c | BD-566610 | 3.9 |
| PE-CF594 | TCR gam-del | BD-562511 | B1 |
| PerCP-Cy5.5 | CD45RO | BD-560607 | UCHL1 |
| Pe-Cy7 | CD56 | BD-557747 | B159 |
| BV605 | CD19 | BioLengend-302244 | HIB19 |
| BV786 | CD16 | BD-563690 | 3G8 |
| BV711 | CD8 | BD-563677 | RPA-T8 |
| BUV395 | CD3 | BD-563546 | UCHT1 |
| BUV496 | CD4 | BD-612936 | SK3 |
| BUV737 | CD14 | BD-564444 | M5E2 |

Supplemental Table 4: Lymphocyte-oriented Panel (S3)

| Fluorochrome | Antigen | Manufacturer/Catalogue # | Clone |
|---|---|---|---|
| FITC | CD38 | BD-560982 | HIT2 |
| PE | CD5 | BD-555353 | UCHT2 |
| APC | CD27 | BD-558664 | M-T271 |
| v450 | CD19 | BD-560353 | HIB19 |
| AF700 | CD138 | Biolegend-356512 | MI15 |
| PerCP-Cy5.5 | CD24 | BD-561647 | ML5 |
| Pe-Cy7 | IgD | BD-561314 | IA6-2 |
| BV605 | PD-1 | Biolegend-329924 | EH12.2H7 |
| BV786 | ALCAM | BD-564939 | 3A6 |
| BV711 | CD8 | BD-563677 | RPA-T8 |
| BUV395 | CD3 | BD-563546 | UCHT1 |
| BUV496 | CD4 | BD-612936 | SK3 |
| BUV737 | CD14 | BD-564444 | M5E2 |

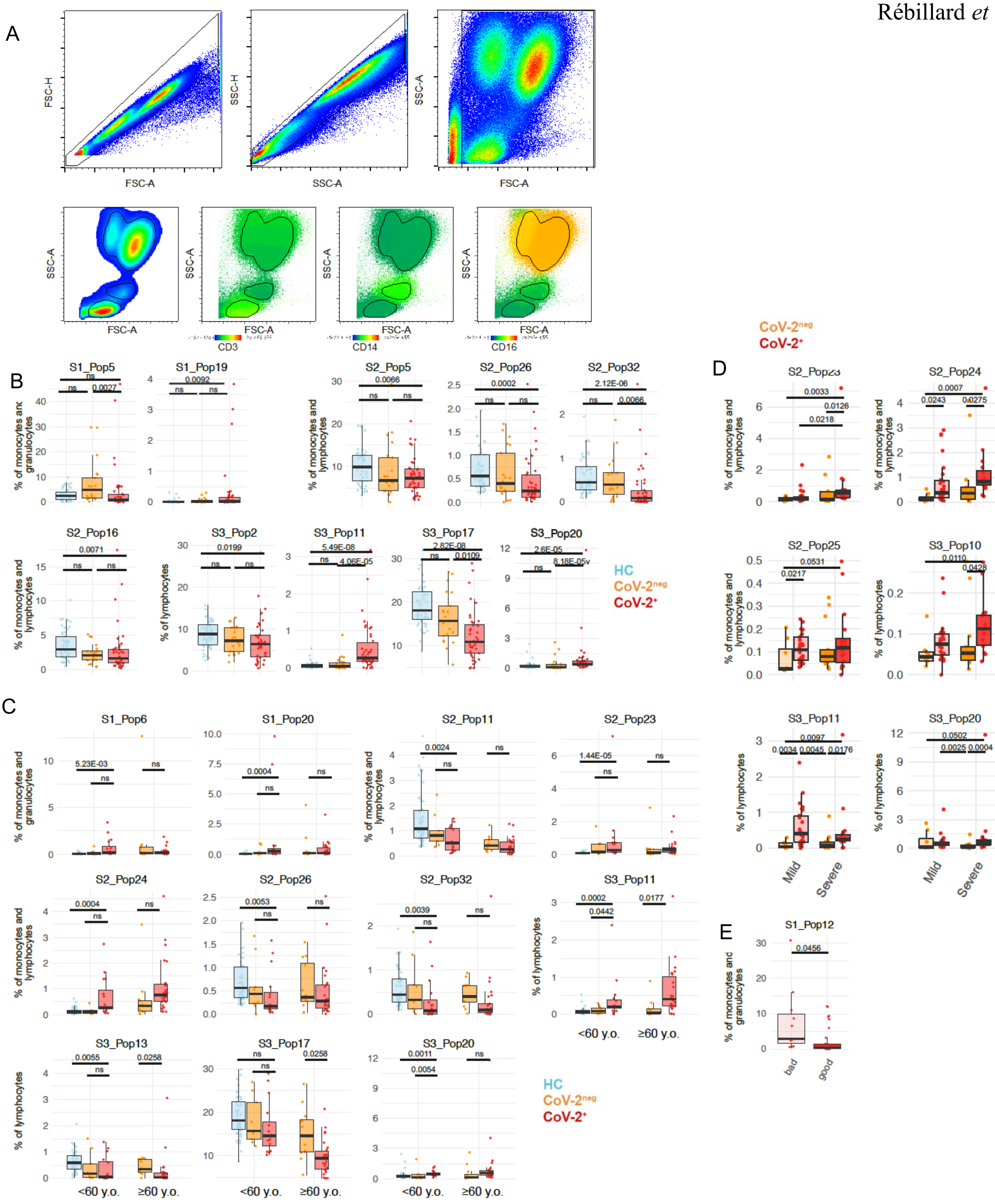


**Supplemental Figure 1. Populations altered in SARS-CoV-2⁺ patients identified by data-driven (unbiased) analysis.** (**A**) Flow cytometry gating strategy for the selection of events that belong to lymphocyte, monocyte/myeloid and granulocyte before the data-driven analysis. (**B-E**) Box and Whisker plots showing frequencies of selected immune cell populations specifically dysregulated in SARS-CoV-$2^+$ patients (CoV-$2^+$); (**B**) CoV-$2^+$ (red) compared to SARS-CoV-$2^{neg}$ (CoV-$2^{neg}$, yellow) and healthy controls (HC, blue) (HC $n$ = 49; CoV-$2^{neg}$ $n$ = 21; CoV-$2^+$ $n$ = 42), (**C**) according to age groups (HC < 60 y.o. $n$ = 49; CoV-$2^{neg}$ hospitalized < 60 y.o. $n$ = 9, ≥ 60 y.o. $n$ = 13; CoV-$2^+$: < 60 y.o. $n$ = 20; ≥ 60 y.o. $n$ = 30), **(D)** according to disease severity (CoV-$2^{neg}$ mild/moderate disease $n$ = 8, severe disease $n$ = 14; CoV-$2^+$ mild/moderate disease $n$ = 29, severe disease $n$ = 21), and **(E)** according to the clinical outcome at 30 days in SARS-CoV-$2^+$ patients (NIH score 5-8, $n$ = 36) vs. (NIH score 1-4, $n$ = 14). Mann-Whitney $U$ test (for $n$ = 2 categories) and Kruskal-Wallis test (for $n$ > 2 categories) followed by a Dunn´s post hoc-test were used. ns: not significant.

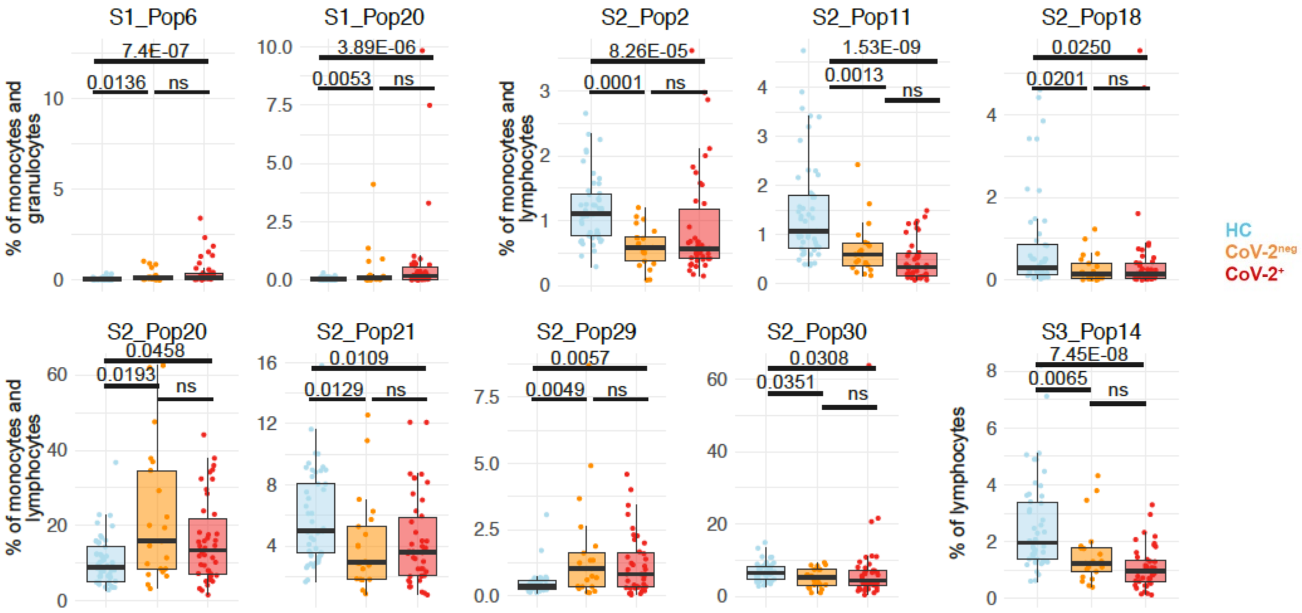


**Supplemental Figure 2. Populations altered in hospitalized patients identified by data-driven (unbiased) analysis**. Box and Whisker plots showing frequencies of selected immune cell populations dysregulated in hospitalized patients compared to healthy controls (HC); SARS-CoV-2$^+$ (CoV-2$^+$, red) compared to SARS-CoV-2$^{neg}$ (CoV-2$^{neg}$, yellow) and HC (blue) (HC $n$ = 49; CoV-2$^{neg}$ $n$ = 21; CoV-2$^+$ $n$ = 42). Mann-Whitney $U$ test (for $n$ = 2 categories) and Kruskal-Wallis test (for $n$ > 2 categories) followed by a Dunn´s post hoc-test were used. ns: not significant.

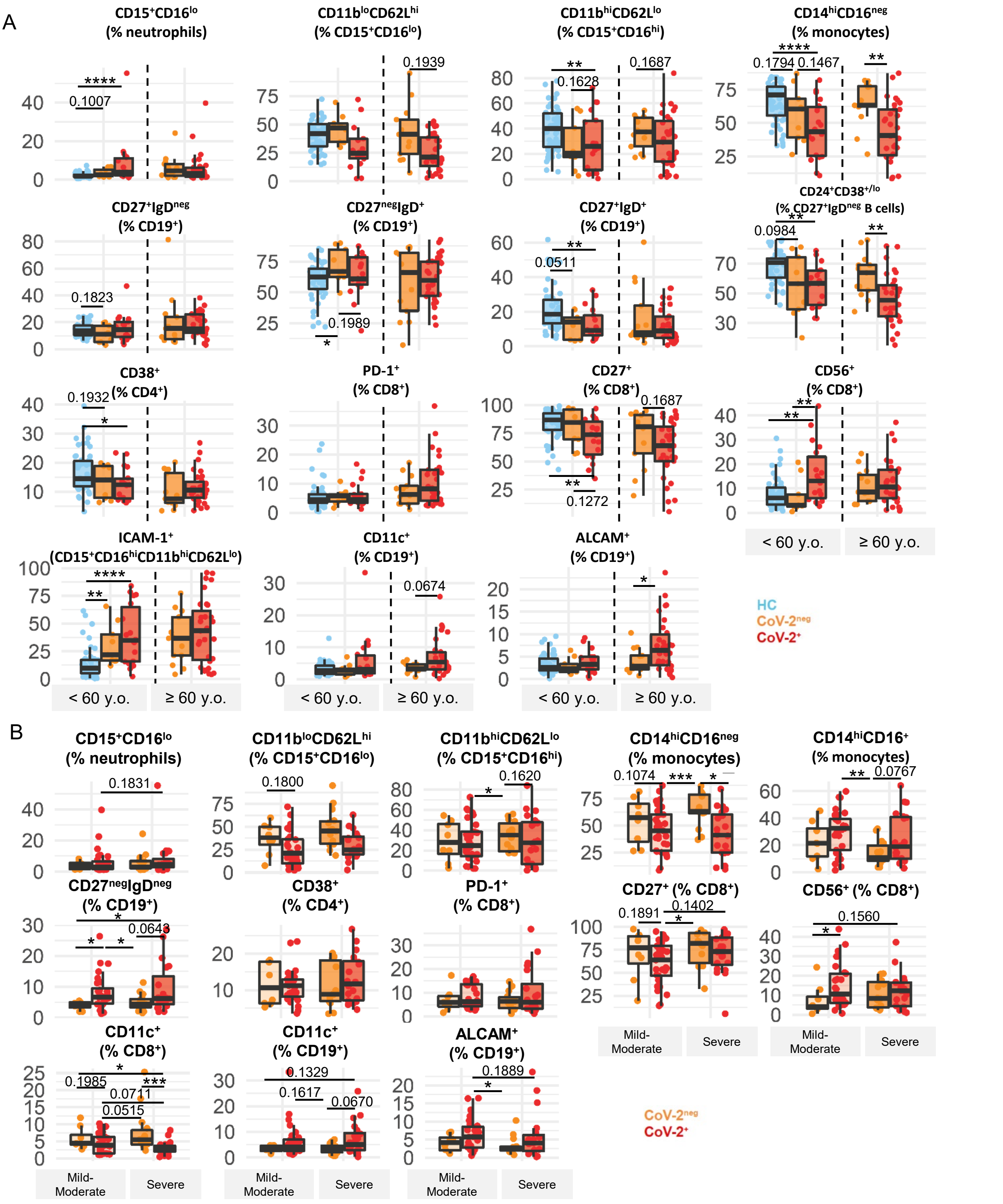


**Supplemental Figure 3. Frequencies of selected immune cell populations identified by conventional analysis.** (**A-B**) Box and Whisker plots showing frequencies of different subsets of immune cell populations in peripheral blood (conventional analysis) from SARS-CoV-2+ (CoV-2+, red) and SARS-CoV-2neg (CoV-2neg, yellow) hospitalized patients and healthy controls (HC, blue) (**A**) according to age groups (HC < 60 y.o. *n* = 49; CoV-2neg hospitalized < 60 y.o. *n* = 9, ≥ 60 y.o. *n* = 13; CoV-2+: < 60 y.o. *n* = 20, ≥ 60 y.o. *n* = 30), and (**B**) according to disease severity in hospitalized patients (CoV-2neg mild/moderate disease *n* = 8, severe disease *n* = 14 ; CoV-2+ mild/moderate disease *n* = 29, severe disease *n* = 21*)*. Mann-Whitney *U* test (for *n* = 2 categories) and Kruskal-Wallis test (for *n* > 2 categories) followed by Dunn's post-hoc test. Each dot represents one donor. *$P$ < 0.05; **$P$ < 0.01; ***$P$ < 0.001; ****$P$ < 0.0001.

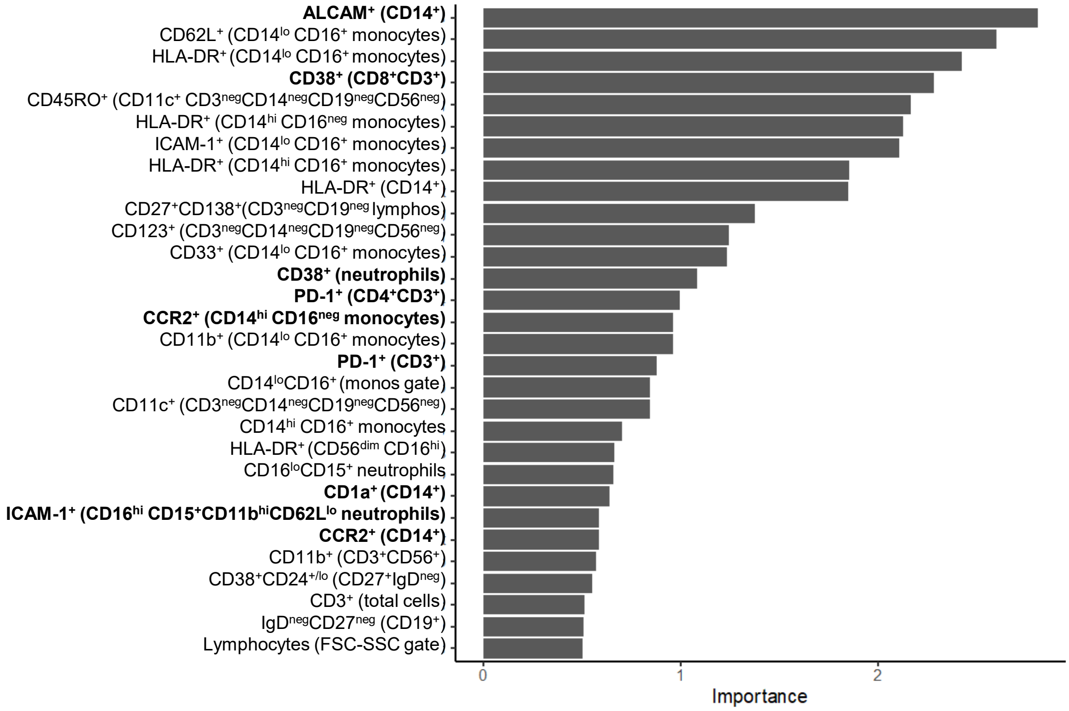


**Supplemental Figure 4. Multivariate model using random forest class prediction analysis.** Results of the top 30 cells population with the greater Gini importance index.
